# Spatio-temporal dynamics of malaria in Zanzibar, 2015–2020

**DOI:** 10.1101/2022.04.26.22274261

**Authors:** Donal Bisanzio, Shabbir Lalji, Faiza B. Abbas, Mohamed H. Ali, Wahida S. Hassan, Humphrey R. Mkali, Abdul-wahid Al-mafazy, Joseph J. Joseph, Ssanyu S. Nyinondi, Chonge Kitojo, Naomi Serbantez, Erik Reaves, Erin Eckert, Jeremiah Ngondi, Richard Reithinger

## Abstract

**Background:** Despite the continued high coverage of malaria interventions, malaria elimination in Zanzibar remains elusive, with the annual number of cases increasing gradually over the last 3 years. The aims of the analyses presented here were to (i) assess the spatio-temporal dynamics of malaria in Zanzibar between 2015 and 2020, and (ii) identify malaria hotspots that would allow Zanzibar to develop an epidemiological stratification for more effective and granular intervention targeting, thereby allowing for more effective programmatic resource allocations.

**Methods:** Data routinely collected by Zanzibar’s Malaria Case Notification (MCN) system were analyzed. The system collects detailed socio-demographic and epidemiological data from all malaria cases who are passively detected and reported at the islands’ 313 public and private health facilities (defined as primary index cases), as well as through case follow-up and reactive case detection (defined as secondary cases). Using these data, spatio-temporal analyses were performed to identify the spatial heterogeneity of case reporting at shehia (ward) level during transmission seasons and its correlation with 2015–2020 rainfall.

**Results:** From January 1, 2015, to April 30, 2020, 22,686 index cases were notified by health facilities and reported through the MCN system. Number of cases reported showed declining trends from 2015 to 2017, followed by an increase from 2018 to 2020. More than 40% of cases had a travel history outside Zanzibar in the month prior to testing positive for malaria. The proportion of followed-up index cases was approximately 70% for all years. Change point analysis identified 10 distinct periods of malaria transmission across the study period, and the seasonality of reported index cases was significantly correlated to the amount of precipitation that occurred during the previous rainy season. Out of Zanzibar’s 388 shehias, 79 (20.3%) were identified as malaria hotspots in any given year between 2015 and 2020; these hotspots reported 52% of all index cases during the study period. Of the 79 shehias identified as hotspots, 12 (3% of all shehias) were hotspots in more than four years, i.e., considered temporally stable, reporting 14.5% of all index cases.

**Conclusions:** Our findings confirm that the scale-up of malaria interventions has greatly reduced malaria transmission in Zanzibar since 2006, with mean annual shehia incidence being 3.8 cases per 1,000 over the 2015–2020 study period. Spatio-temporal analyses identified hotspots, some of which were stable across multiple years. From a programmatic perspective, malaria efforts should progress from an approach that is based on universal coverage of interventions to an approach that is more tailored and nuanced, with resources prioritized and allocated to a select number of hotspot shehias.

## Background

Malaria remains a major global public health concern, with an estimated 241 million malaria cases and 627,000 deaths reported across 87 endemic countries in 2020.[1] From 2003 onwards, the Zanzibar Malaria Elimination Program (ZAMEP) gradually introduced and scaled-up rapid diagnostic tests (RDTs), artemisinin-combination based therapies (ACTs), intensive vector control (long-lasting insecticidal nets [LLINs] and indoor residual spraying [IRS]) and case-based surveillance, all of which resulted in a large decline of malaria cases and deaths. Thus, malaria prevalence in children under 5 years of age decreased from 40% before 2002 to less than 1% in 2007/8, and has remained at those low levels ever since.[2-4] Nonetheless, despite the continued high coverage of malaria interventions, malaria elimination in Zanzibar remains elusive. Thus, from 2015 to 2020, the annual number of reported malaria cases and malaria incidence have gradually increased, the annual number of severe malaria admissions increased from 89 to 606, and the annual number of malaria-related deaths increased from 1 to 20.[5] The reasons for this resurgence are likely due to a combination of factors, including residual transmission with more outdoor rather than indoor biting mosquito vectors;[6] asymptomatic (and undetected) malaria infections in the community that remain a possible source for local transmission;[7] insecticide resistance in mosquito vectors that affect the efficacy of LLINs and IRS; and a variability in environmental factors important for mosquito and parasite reproduction and survival.[4, 8]

As countries progress toward malaria elimination, strengthening malaria surveillance systems and using the data collected through these systems to better understand malaria transmission dynamics at a more granular spatio-temporal scale becomes increasingly important.[9] Spatio-temporal heterogeneity in transmission dynamics is likely to occur in elimination settings, with certain locations having no or very little transmission, and other locations (so-called “hotspots”) experiencing comparatively high transmission. Stresman et al. [10] defined malaria hotspots as areas “where transmission intensity exceeds the average level”, adding that hotspots are, typically, <1 km^2^ in size and are often within a focus of active malaria transmission. Furthermore, they argued that a hotspot that is conducive of transmission across both dry and rainy seasons is stable (rather than unstable), and thus should be able to be detected across time. It is assumed that these hotspots disproportionally contribute to maintaining ongoing transmission and that targeting them will achieve greater impact as well as maximize available resource allocation. Consequently, identifying hotspots and characterizing whether these hotspots persist over time may help in stratifying Zanzibar epidemiologically, to then determine the level and type of interventions, as well as the amount of programmatic and financial resources that are needed to progress towards malaria elimination.

The aims of the analyses presented here were (i) to describe the spatio-temporal dynamics of malaria in Zanzibar between 2015 and 2020, and (ii) identify malaria hotspots that would allow ZAMEP and stakeholders to develop an epidemiological stratification for more effective and granular targeting of malaria interventions, thereby maximizing programmatic resource allocations.

## Methods

### Study setting

The archipelago of Zanzibar is located between longitudes 39.19793 and latitudes -6.16394, 25–50 kilometers off the east coast of the Tanzania mainland in the Indian Ocean. There are two main islands, Pemba and Unguja, which cover a total land area of 2,461 km^2^ and have an estimated population of 1,717,608 people. Zanzibar comprises 11 districts, which are subdivided into 387 shehias, 258 of which are on Unguja and 129 are on Pemba. Shehias are akin to wards and Zanzibar’s lowest administrative unit, where many of the public services are planned, managed, and implemented, including for health and malaria.

Zanzibar’s climate is characterized by two main rainfall seasons: a primary (March–May, called *masika*) and a secondary (November–January, called *vuli*) season; rainfall is at its lowest in July. Both rainfall seasons are followed by peak malaria transmission seasons, with the highest malaria case count typically observed in the March–May rainfall season.

### Data collection

Data used in this study had been routinely collected by ZAMEP’s Malaria Case Notification (MCN) system between January 1, 2015, and April 30, 2020. The MCN system was established in 2012 to electronically collect detailed socio-demographic and epidemiological data from all malaria cases in Zanzibar in order to inform programmatic decision-making—both for cases passively diagnosed by microscopy or rapid diagnostic tests (RDTs) at health facility level (defined as primary index cases), as well as cases diagnosed by RDTs during case follow-up and reactive case detection (rACD) activities at household level (defined as secondary cases). By 2014, the system had been progressively scaled-up to cover all 189 public and 124 private health facilities on Pemba and Unguja.

Suspected malaria cases access health facilities, where they get tested for malaria. If confirmed as positive, the health provider prescribes ACTs as per national malaria diagnosis and treatment guidelines.[11] Within 24 hrs of the index case being detected at facility-level, the provider sends an unstructured supplementary service data (USSD) notification to a central ZAMEP computing server. The notification is forwarded to a District Malaria Surveillance Officer (DMSO), who visits the health facility to confirm the reported index case and collects additional information, including the patient’s contact details. Within 48 hrs of being notified, the DMSO then follows-up index cases at household-level, ensuring they are adhering to prescribed treatment and investigating case details that will inform case classification (e.g., whether the case was autochthonous versus possibly imported because of a travel history in the preceding 30 days). DMSOs then use RDTs (in most cases and years the SD Bioline® HRP2 / pLDH RDT from Standard Diagnostics, Giheung-ku, Republic of Korea, was used) to screen all of the index case’s additional household members; members with a positive RDT result (i.e., secondary cases) are treated with an ACT. Using an electronic, standardized questionnaire that is completed by the DMSO, the case follow-up and reactive case detection (rACD) data, including for household-based screening and treatment (HSaT), are linked to each index case through the Coconut surveillance platform (https://coconutsurveillance.org/). The specific variables collected in the questionnaire are individual factors (i.e., contact information, age, sex, self-reported history of travel in the last 30 days, self-reported history of fever in the last two weeks, RDT positivity, and LLIN use the previous night); household factors (i.e., number of people residing in the household, number of household LLINs, and IRS application in the last 12 months); and geographical factors (i.e., household geolocation and weekly rainfall). During the 2015–2020 study period, rainfall data were obtained from 10 meteorological stations managed by Zanzibar’s Tanzania Meteorological Agency, with rainfall data measured in millimeters (mm) and recorded daily.

### Data analysis

In Zanzibar, malaria transmission occurs throughout the year, and it is characterized by two high-transmission periods after the vuli and masika rainy seasons. The duration of each high-transmission season, as well as the following low-transmission season, varies among the years due to precipitation patterns. To identify the duration of each transmission season during the 2015–2020 study period, we used the change point analysis technique to analyze the weekly trend of malaria cases.[12]. This technique enables to divide times series by identifying those points in time (i.e., change points) when substantial changes occur in a data trend. The change points identified by the analysis were used to estimate the duration of each malaria transmission season throughout any given study year.

The spatial pattern of malaria index cases during each transmission season from 2015 to 2020 was assessed using a hotspot analysis based on the Getis-Ord G_i_* local spatial clustering test.[13] The results of the G_i_* local spatial test were used to identify hotspot shehias; shehia was used as the geographic unit to define hotspots, since it is the lowest administrative unit at which programmatic decision-making occurs. The significance of the computed G_i_* was estimated by comparing observed values to the random case distribution (null hypothesis) by randomly re-assigning the weekly cases to the shehias. The statistical significance calculation was based on 100,000 Monte Carlo randomizations (p<0.05, with Bonferroni correction). A Kendall’s concordance coefficient (W) was calculated to investigate the spatio-temporal overlap of hotspots from 2015 to 2020.[14] Kendall’s W measures concordance between two datasets, ranging from +1 (complete agreement) to -1 (no agreement). The relationship between reported cases and rainfall was evaluated using the auto-correlation function.[15]

### Definition of indicators

Based on the G_i_*, hotspot shehias were defined as shehias with a statistically significant higher number of malaria index cases compared to their neighboring shehias in any given transmission season. We defined hotspot shehias as temporally stable, if during the five-year study period, they were classified as hotspots either during the high or low transmission season for at least 4 out of the five study years. The probability of finding a positive secondary case (detection rate) was defined as the number of secondary cases detected through case follow-up and rACD of each primary index case. The probability to find a secondary case and its 95% confidence interval (95% CI) was estimated using a logistic regression.

### Ethical approval

Ethical clearance to undertake secondary analysis of the MCN surveillance data was granted by the Zanzibar Health Research Institute (ZAHRI) with reference number ZAHREC/03/AUG/2021/20. All personal identifiers were removed during data cleaning before analysis.

## Results

### Index Cases

From January 1, 2015, to April 30, 2020, 27,046 cases were notified by health facilities in Zanzibar, of which 22,686 were followed-up at health facility level by DMSOs and reported through the MCN system (called [DMSO] index cases). The number of these index cases showed a declining trend from 2015 to 2017, followed by an increase from 2018 to 2020 (Table 1; Figure 1 and Figure 2).

**Table 1.** Summary of reported index cases from January 1, 2015, to April 30, 2020, in Zanzibar.

|  | 2015 | 2016 | 2017 | 2018 | 2019 | 2020<sup>a</sup> |
| --- | --- | --- | --- | --- | --- | --- |
| Cases notified by health facilities | 4,325 | 3,856 | 4,252 | 5,494 | 6,766 | 6,678 |
| Number of index cases followed up to health facilities by DMSOs | 3,745 | 2,596 | 2,940 | 3,647 | 5,747 | 4,011 |
| Number of investigated index cases (%) | 2,592<sup>b</sup><br>(69.2%) | 2096<br>(79.7%) | 2081<br>(70.8%) | 2736<br>(75.0%) | 4259<br>(74.1%) | 2,630<br>(65.6%) |
| Median age (years) of investigated index cases | 20<br>(IQR: 11–29) | 19<br>(IQR: 9–28) | 21<br>(IQR: 13–29) | 20<br>(IQR: 11–29) | 22<br>(IQR: 13–31) | 23<br>(IQR: 15–31) |
| Fraction of investigated index cases who were male | 59.3% | 57.6% | 62.6% | 59.3% | 64.8% | 69.8% |
| Fraction of investigated index cases reported in Unguja | 81.6% | 69.7% | 84% | 77.1% | 82.9% | 80.2% |
| Number of index cases with travel history outside Zanzibar (%) | 1,926<br>(51.4%) | 1,195<br>(46.0%) | 1,223<br>(41.6%) | 1,938<br>(53.1%) | 3,536<br>(61.5%) | 1,365<br>(34.0%) |
| Number of index cases with travel history within Zanzibar (%) | 139<br>(3.7%) | 87<br>(3.4%) | 87<br>(3.0%) | 154<br>(4.2%) | 197<br>(3.4%) | 220<br>(5.5%) |
| Fraction of index cases reported during January–April who have travel history outside Zanzibar | 62.7% | 51.0% | 55.2% | 58.6% | 69.0% | 35.1% |
| <sup>c</sup> Fraction of index cases reported during January–April who have travel history within Zanzibar | 4.4% | 4.9% | 3.0% | 4.6% | 3.0% | 5.7% |
| Weekly rainfall year in mm (median [IQR]) | 9.8<br>(1.9–30.7) | 2.9<br>(0.0–18.3) | 15.5<br>(3.1–72.6) | 21.8<br>(6.9–63.5) | 16.6<br>(2.4–88.9) | 10.9<br>(6.0–12.9) |
| Weekly rainfall during the masika season in mm, mid-March–May (median [IQR]) | 52.4<br>(12.5–96.3) | 19.8<br>(3.7–55.1) | 125.6<br>(62.7–175.7) | 56.3<br>(25.2–129.3) | 43.3<br>(13.3–84.6) | – |
| Weekly rainfall during the vuli season in mm, November–mid January (median [IQR]) | 11.8<br>(4.5–43.8) | 13.3<br>(2.6–19.3) | 19.2<br>(9.1–66.3) | 17.9<br>(9.9–24.8) | 43.8<br>(15.7–74.4) | – |

<sup>a</sup>Time period January 1 to April 30; <sup>b</sup>data available from January 1 to September 16; <sup>c</sup>Calculated to make comparison among all years from 2015 to 2020

**Table 2.** Summary of reported secondary cases from January 1, 2015, to April 30, 2020, in Zanzibar.

|  | 2015 | 2016 | 2017 | 2018 | 2019 | 2020<sup>a</sup> |
| --- | --- | --- | --- | --- | --- | --- |
| Number of household members tested during index case investigation | 11,601<sup>b</sup> | 8,479 | 7,638 | 10,782 | 14,963 | 8,511 |
| Median age (years) of investigated index cases | - | 15<br>(IQR: 7–25) | 16<br>(IQR: 8–26) | 16<br>(IQR: 7–25) | 18<br>(IQR: 8–28) | 17<br>(IQR: 8–26) |
| Fraction of index cases who are male | 49.7% | 41.6% | 43.7% | 47.4% | 48.5% | 53.0% |
| Number of positive investigated people (secondary cases) (%) | 591<sup>b</sup><br>(5.1%) | 365<br>(4.3%) | 319<br>(4.2%) | 445<br>(4.1%) | 544<br>(5.8%) | 267<br>(3.7%) |
| Number of positive investigated people (secondary cases) performed from January–April (%) | 169<br>(6.7%) | 167<br>(4.9%) | 74<br>(4.7%) | 164<br>(4.8%) | 177<br>(5.1%) | 267<br>(3.7%) |
| Probability to find a secondary case during investigation (95% CI) | 0.048<sup>b</sup><br>(0.045–0.052) | 0.041<br>(0.037–0.045) | 0.040<br>(0.036–0.044) | 0.040<br>(0.036–0.043) | 0.035<br>(0.032–0.038) | 0.030<br>(0.027–0.034) |

**Figure 1.**
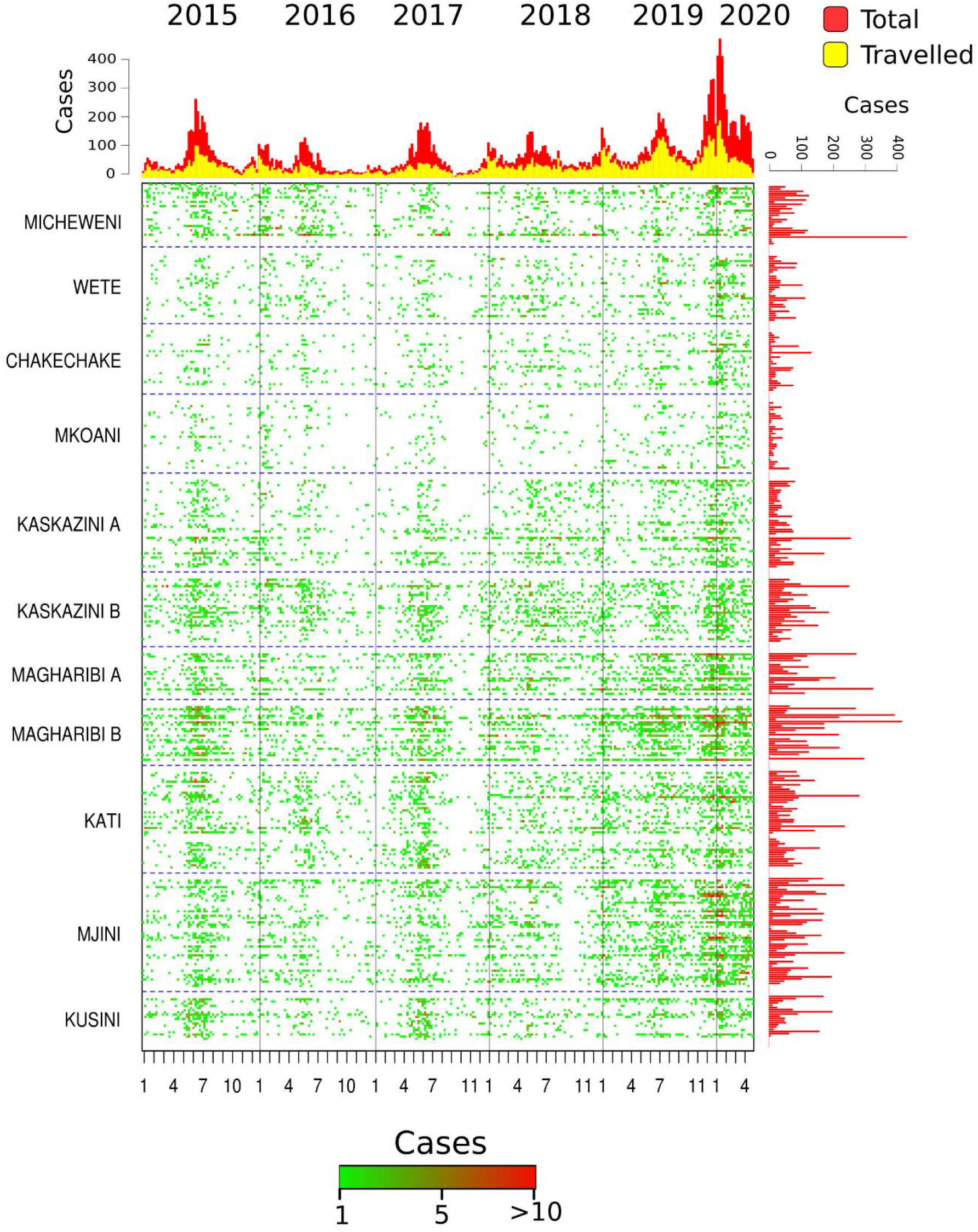
Number of confirmed malaria cases per shehia from January 1, 2015, to April 30, 2020. Each row of the plot represents the time series of reported index cases per week per shehia. The shehias were grouped by district ordered by latitude (from North to South). The image shows the cumulative number of index cases per year split by travel history (Top; horizontal) and cumulative number of cases per shehia during the 2015–2020 study period (Right; vertical).

**Figure 2.**
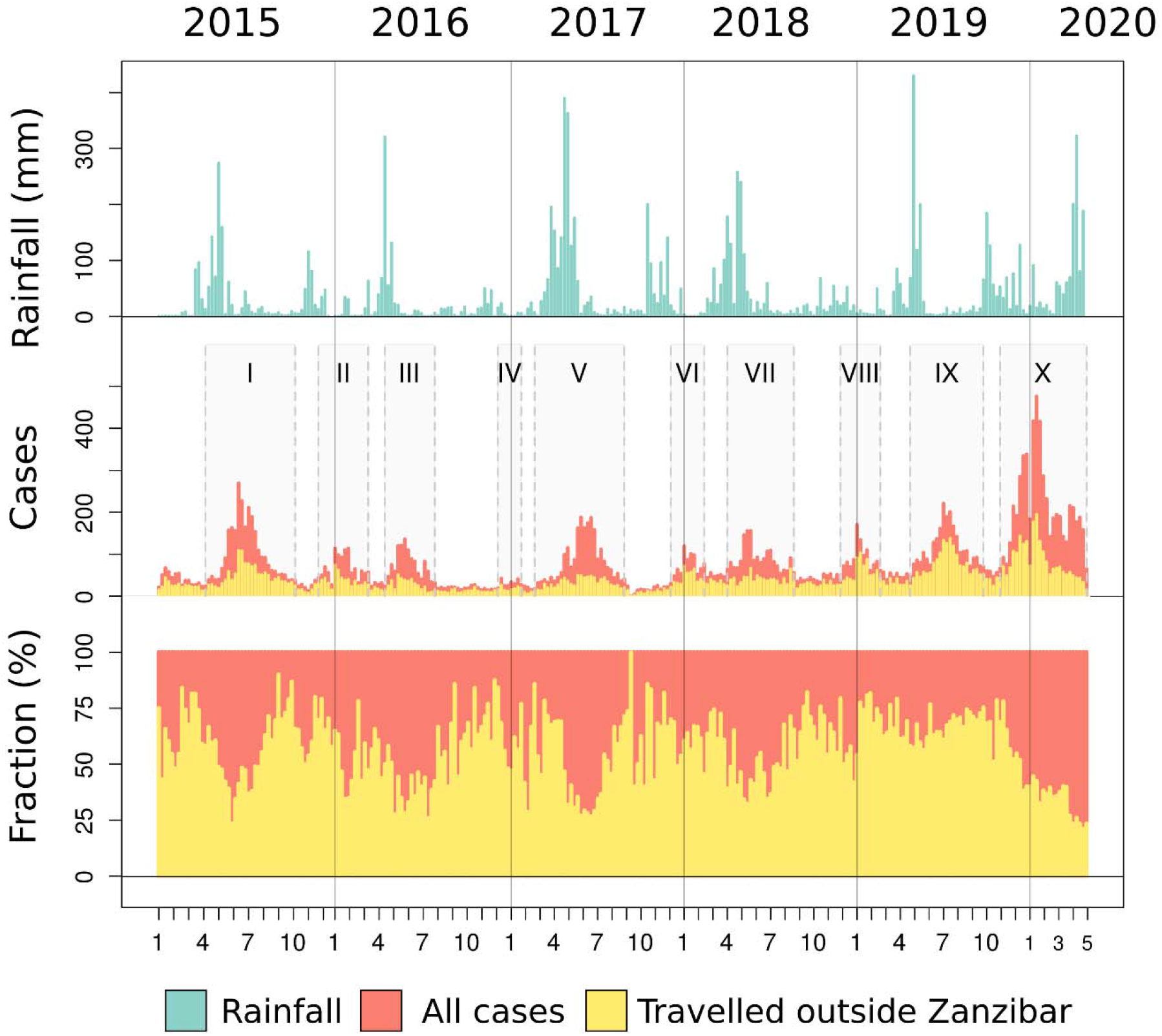
Weekly rainfall and reported malaria index cases in Zanzibar from January 1, 2015, to April 30, 2020. The figure also shows the number and fraction of index cases with travel history outside Zanzibar (yellow bars). The grey boxes indicate the ten high transmission seasons of the study period; transmission seasons were classified using Roman numerals.

Median age of index cases was 21 (interquartile range [IQR]: 12–30) years, with 62.8% of cases being male; neither age nor sex ratio significantly varied across study years. Of index cases, 79.9% were reported in Unguja (range across years: 69.7–84.0%) (Table 1). Comparing index cases reported from January 1 to April 30 of each year from 2015 to 2020, 2019 showed the highest number of reported cases. The proportion of cases that was followed-up to household level was approximately 70% for all years, with 2016 being highest (79.7%) (Table 1).

Among reported index cases over the entire 2015–2020 study period, more than 40% had a travel history outside of Zanzibar in the month prior to testing positive for malaria. The proportion of cases who reported traveling outside of Zanzibar prior to confirmatory testing decreased from 2015 to 2017, increased in 2018 and 2019, before decreasing again in 2020.

### Secondary Cases

The percentage of tested household members who were malaria positive (secondary cases) showed a declining trend from 2015 to 2018, before increasing in 2019 (Table 1). Comparing index cases reported from January 1 to April 30 of each year, 2020 showed the lowest number of tested household members who were malaria positive. The probability of finding a positive person (detection rate) after index case investigation was approximately 0.04 (i.e., 1 additional secondary case was detected for every 400 investigated index cases) and was significantly higher during 2015 compared with the other years. The trend of the detection rate slightly declined from 0.041 in 2016 to 0.030 in 2020.

### Malaria Transmission Seasons and Precipitation

Change point analysis identified 10 distinct periods of malaria transmission across the study period (Figure 2). Although these generally aligned with the expected peak transmission seasons following the masika and vuli rainy seasons, the analysis showed how variable the seasons were across years, both in terms of onset as well as duration. From 2015 to 2018, precipitation during the vuli season tended to be lower compared to the masika rainy season. From 2017 to 2020, precipitation increased in both rainy seasons, and in 2019 precipitation during the vuli and masika rainy seasons were similar (Table 1). The seasonality of reported index cases was significantly correlated to the amount of precipitation that occurred during the previous rainy season (Figure 2 and Figure 3). Cross-correlation analyses showed that the number of index cases had the highest correlations with total precipitation in the 12th and 13th weeks prior to malaria case confirmation (Figure 3).

**Figure 3.**
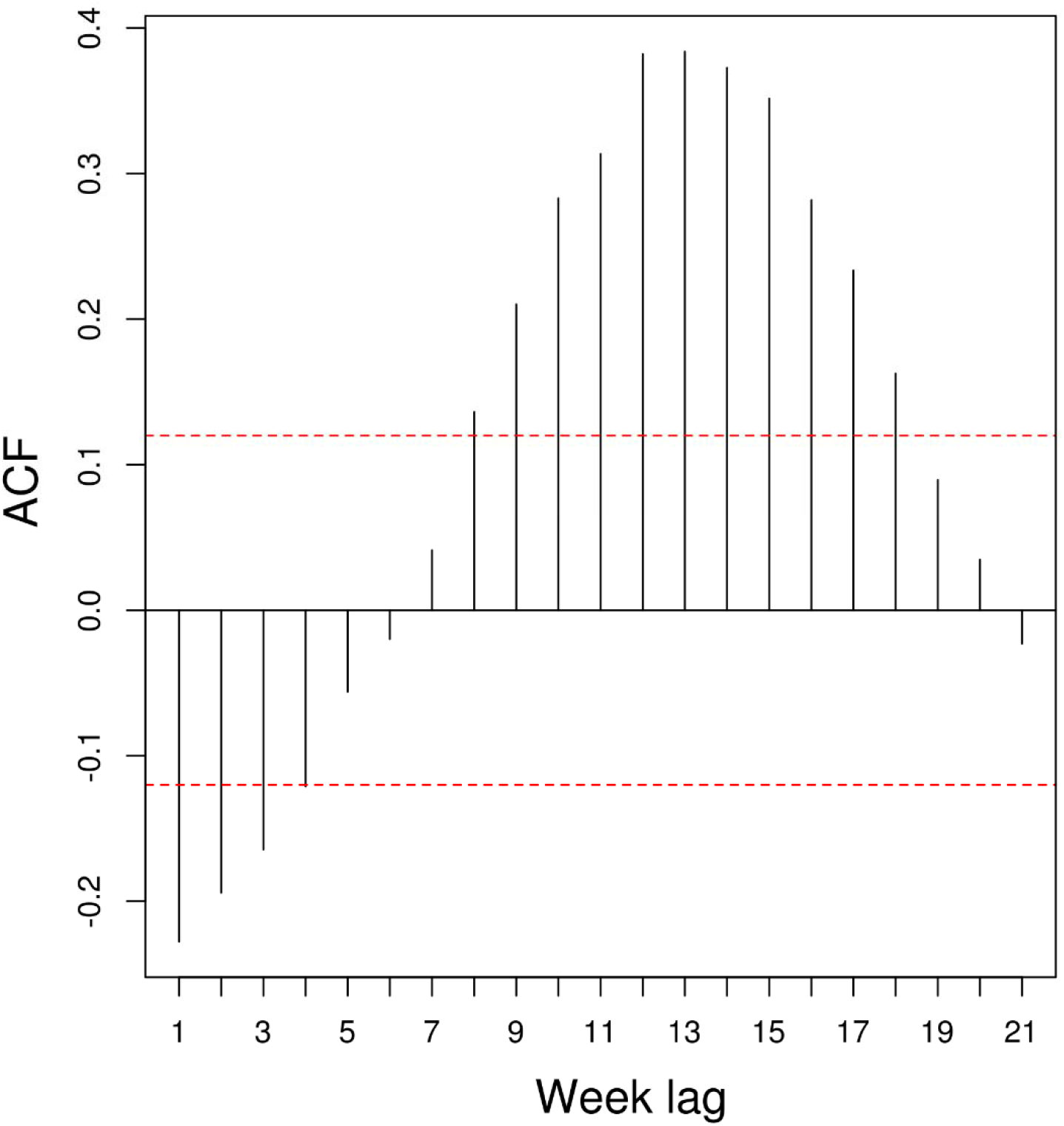
Cross-correlation of weekly reported index cases and weekly precipitation. The black lines passing the red dotted lines are significant correlations, p<0.05.

### Spatio-temporal Dynamics and Identification of Hotspots

During peak transmission seasons, Unguja reported more index cases than Pemba (Figures 1, 4, and 5). Additionally, the spatial pattern of the reported index cases in the two islands was different. Most of the index cases reported on Unguja were from shehias in the southern part of the island (Figures 1, 4, and 5). On Pemba, northern shehias reported more cases compared to the rest of the island (Figures 1, 4, and 5). Of all 387 shehias, 54 (14.0%) did not report any index cases during the study period.

**Figure 4.**
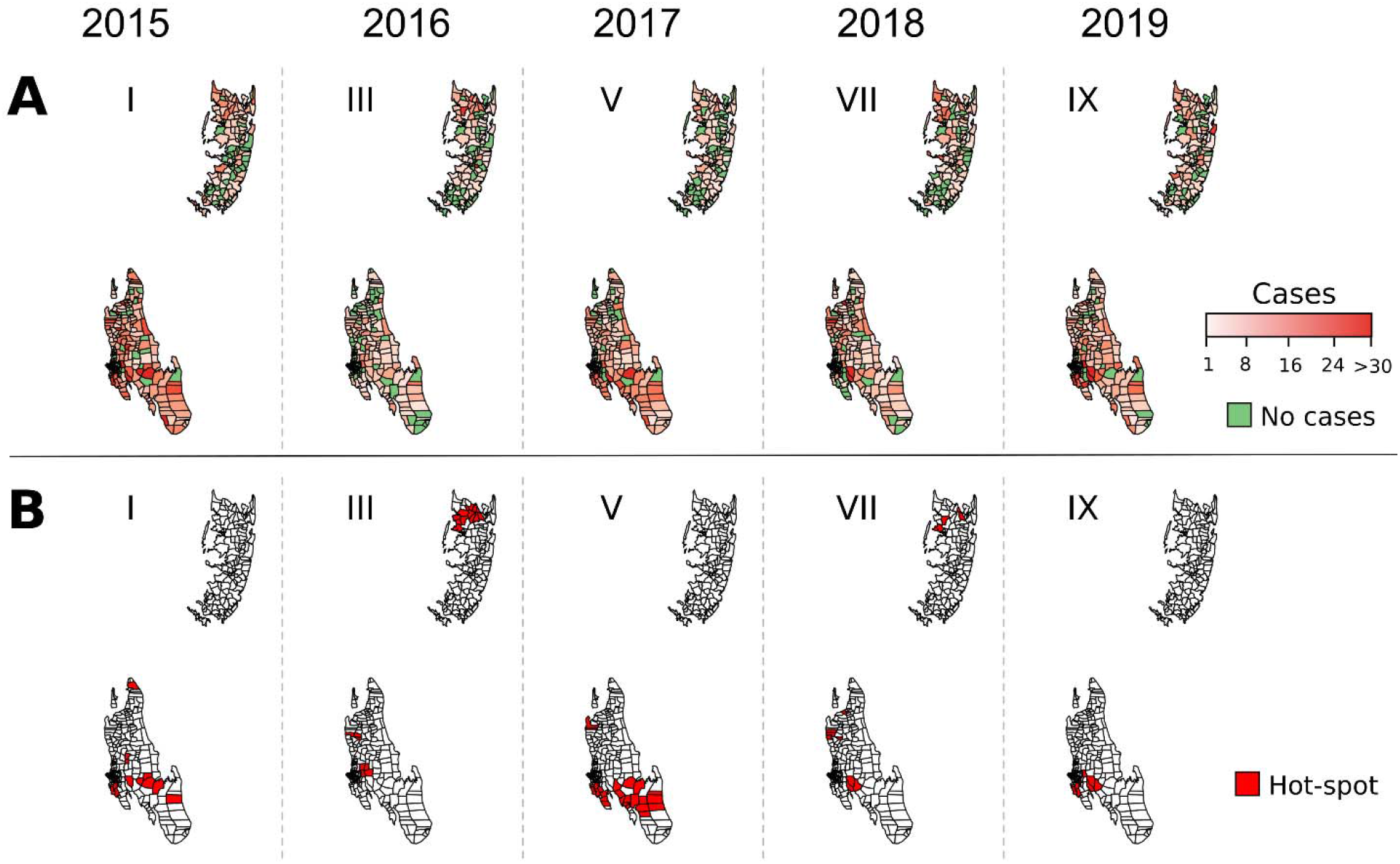
Number of malaria index cases per shehia (A) and hotspot shehias (B) during the high transmission seasons after the masika season from January 1, 2015, to April 30, 2020. The Roman numerals are the various transmission seasons as specified in Figure 2. Hotspots were considered significant when p<0.05 as per G_i_* local spatial clustering test.

**Figure 5.**
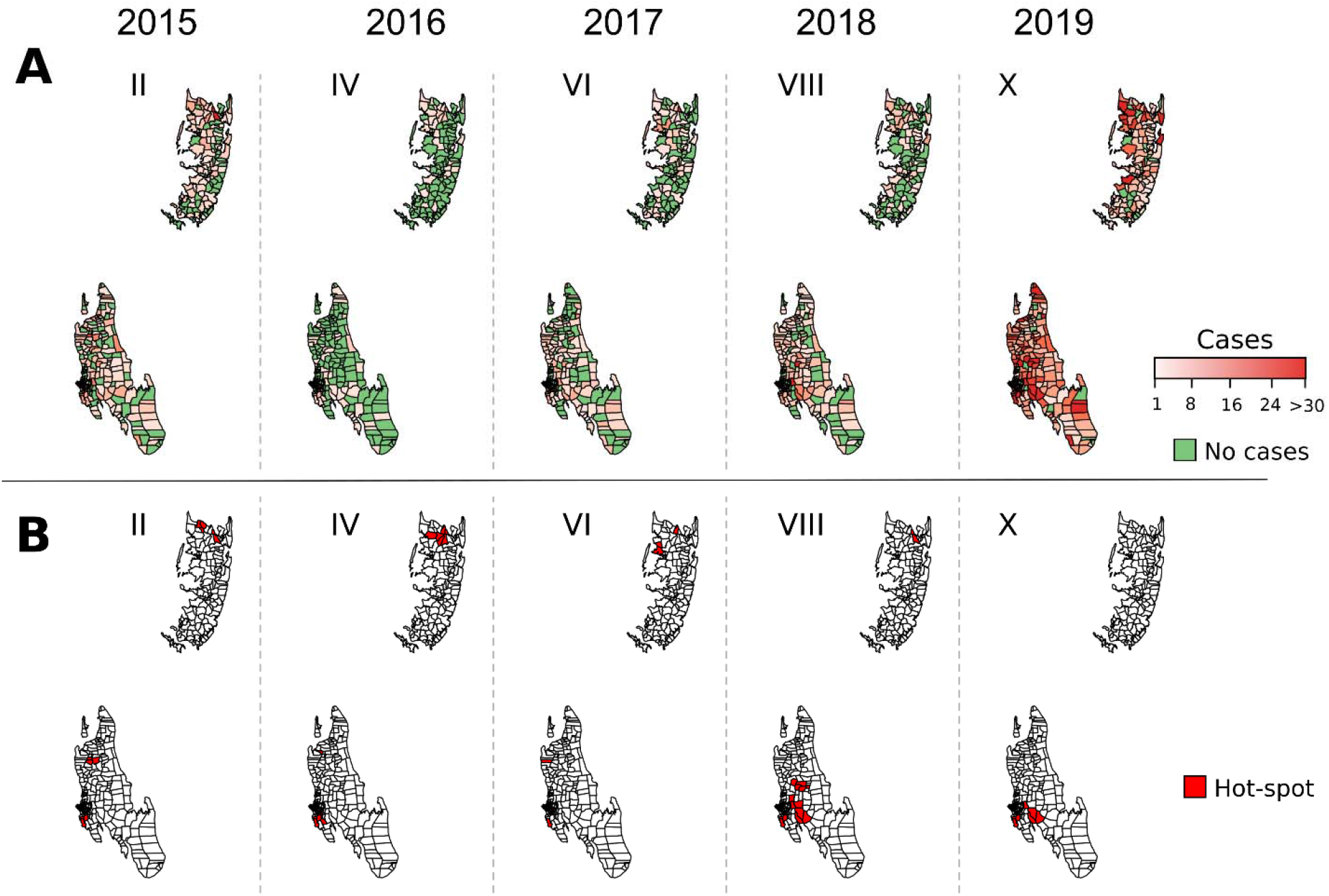
Number of malaria index cases per shehia (A) and hotspot shehias (B) report during high transmission seasons following the vuli season from January 1, 2015, to April 30, 2020. The Roman numerals are the various transmission seasons as specified in Figure 2. Hotspots were considered significant when p<0.05 as per G_i_* local spatial clustering test.

The results from the analyses identified shehia hotspots in the north-eastern and the southern part of Unguja during the 2015–2020 study period; hotspot shehias on Pemba were located in the northern part of the island (Figures 4, 5, and 6). In 2019 and 2020, the spatial pattern of index cases reported during the peak transmission season after the masika and vuli rainy seasons (i.e., transmission seasons IX and X) were similar (Figures 4 and 5). In all other years, the transmission season after the masika rainy season was longer compared to the transmission seasons that occurred after the vuli rainy season (Figures 4 and 5).

**Figure 6.**
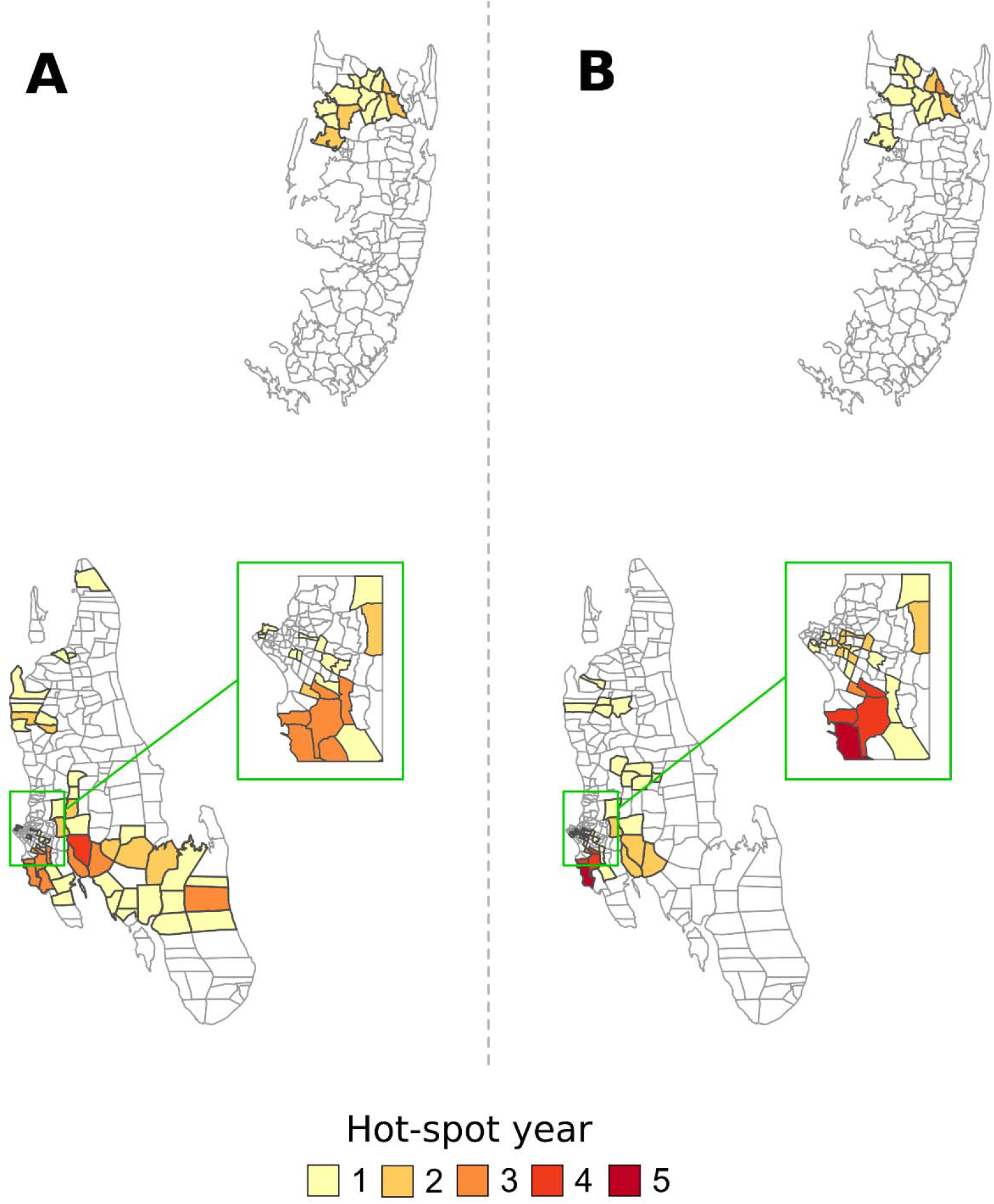
Number of years in which each shehia was identified as hotspot during peak transmission seasons following the masika (A) and vuli (B) rainy seasons from January 1, 2015, to April 30, 2020. Non-highlighted shehias were not identified as hotspot during the study period as per local Moran’s I.

Across Unguja and Pemba shehias, 64 (24.8%) and 15 (11.6%) shehias were identified as a hotspot in any given year, respectively, i.e., reported malaria index cases in either transmission season were observed to significantly cluster spatially and temporally in those shehias. When the hotspot shehias’ temporal stability was examined, it varied greatly between years. Of hotspot shehias, 12 (3.1%) were temporally stable, i.e., they were identified as hotpots either during the high or low transmission season for at least 4 out of the five study years (Figure 6). The 12 hotspots shehias that were stable reported 3,294 (14.5%) of all reported index cases during the study period. Conversely, the 67 shehias that were identified as non-stable hotpots—that is, they were identified as hotpots for <4 years during 2015–2020—contributed to 37.5% (8,519 cases) of reported index cases during the study period.

## Discussion

Our analyses confirm the low level of malaria burden in Zanzibar,[2–4] with annual shehia incidence ranging between 0–54.5 cases per 1,000 population (mean equal to 3.8 cases per 1,000) over the 2015– 2020 study period. Most of the cases in Zanzibar are passively detected at public or private facilities, with cases detected through rACD being comparatively low: over the entire study period, the proportion of all cases that were reported through rACD ranged between 6.5% and 29.4% between years, and the probability to detect such a secondary case during rACD ranged from 0.030 to 0.048. We also confirm previous analyses that show a significant correlation between monthly rainfall and confirmed malaria diagnosis,[2, 3, 8] as well as that a large proportion of cases (34.0–61.5% for the entire study period; up to 69.0% in 2019, if reporting is limited to the January–April peak transmission period) have a reported history of travel outside of Zanzibar one month prior to testing positive for malaria infection.[16]

Studies analyzing the spatio-temporal distribution of malaria have traditionally relied on surveys of well-defined at-risk populations,[17–21] but increasingly—as countries’ health management information systems have been strengthened—studies have used routine PCD [17, 18, 20–25] to define clusters of malaria risk. These studies reported variable patterns of spatio-temporal clustering. Thus, some studies reported the existence of consistent hotspots [24, 26], while others suggest greater variability [17, 18, 20, 21]. For example, in Ouagadougou, Burkina Faso, the location of clusters identified as high risk varied little across three transmission periods [24]. In contrast, in Kilifi county, Kenya, only two temporally stable hotspots were identified over the 1-year study period, comprising 2.7% of all study households and contributing to 10.8% of all malaria cases confirmed by RDT.[27]

Using PCD, our findings support the hypothesis that—in an elimination setting such as Zanzibar—malaria tends to significantly cluster within certain hotspot geographic units.[17, 20, 21, 28, 29, 30] Across Zanzibar’s shehias, 79 (20.4%) were identified as a hotspot in any given year, with malaria observed to significantly cluster spatially and temporally. These hotspot shehias contributed disproportionally to the number of reported malaria index cases, with 52% of all index cases during the study period being reported from there. Similarly, in the 12 stable hotspot shehias (i.e., 3% of all shehias), 14.5% of all index cases were reported.

The use of routinely collected case data through the MCN system does offer the opportunity to detect malaria case clusters down to the shehia, village and household levels at an affordable cost. Interventions tailored and targeted to hotspots have been hypothesized to be highly efficient method in reducing malaria transmission not only inside these hotspots, but also in adjacent geographic areas.[10] While biologically plausible, so far there has been mixed evidence to support this concept. For example, in Rufiji District on mainland Tanzania, locally tailored and targeted interventions contributed to reduce malaria transmission in hotspot villages.[25] Similarly, on Sabang island, Indonesia, an intensified application of malaria diagnosis, ACTs, LLINs and IRS in hotspot areas contributed to a 30-fold reduction in malaria incidence from 3.18 to 0.13 per 1,000 population.[31] In contrast, a trial in western Kenya targeting hotspots with intensified interventions—larviciding, LLINs, IRS and mass drug administration—failed to result in any sustained reduction in malaria transmission in targeted hotspots and failed to impact malaria transmission outside of targeted areas.[32]

For Zanzibar, a number of more aggressive programmatic approaches to reduce malaria transmission in hotspots shehias could be envisaged, including screen and treat strategies, potentially using a more highly sensitive diagnostic test,[33–35] or targeted mass/focal drug administration.[36, 37] Since travel-related malaria represents a large proportion of detected cases, additional interventions targeting travelers, especially prior to high transmission seasons should be considered, including chemoprophylaxis for anyone traveling from Zanzibar to mainland Tanzania, and mass screening and treatment or presumptive treatment of anyone arriving from the mainland to Zanzibar.[9, 38–40]

### Limitations

A number of potential caveats of our analyses should be highlighted, most of which are due to the fact that we used routinely collected programmatic data from ZAMEP in our analyses and not data from a carefully controlled academic research study. First, malaria testing to identify cases is RDT-based, which in the context of Zanzibar is known to have—depending on infections’ parasite densities—low to moderate sensitivity;[41, 42] consequently, it is likely that low parasitemia, asymptomatic infections were missed. It is unlikely, however, that such infections would have clustered in a specific spatio-temporal pattern that is different from the findings we report here, and, thus, would unlikely change our findings and conclusions. Second, depending on the week, month and year, a range (e.g., between 65.6% and 79.7% for any given year) of index cases were followed-up and investigated. Most often such variability in case follow-up and investigation is due to DMSO bandwidth availability resulting from a high incidence of malaria cases—the more index cases are detected and reported at health facilities, the more probable it is that DMSO will not be able to follow-up all index cases and visit their households. Third, we delineated our hotspots to the shehia boundaries, rather than a defined area size (e.g., 1 km^2^). This was done because shehias are the lowest administrative unit that plan, implement, and monitor malaria programming in Zanzibar; any response to an increase in cases would occur at shehia or—depending on the size of the increase—district level.

## Conclusions

The scale-up of malaria interventions has greatly reduced malaria transmission in Zanzibar since 2006, with mean annual shehia incidence being 3.8 cases per 1,000 over the 2015–2020 study period. In our analyses, we identified 79 (20.3%) of Zanzibar’s shehias as malaria hotspots in any given year between 2015 and 2020; 12 of these shehias were considered temporally stable.

From a programmatic perspective, we recommend that malaria efforts should progress from an approach that is based on universal coverage of interventions to an approach that is more tailored and nuanced, with resources prioritized and allocated to a select number of geographic units, i.e., hotspot shehias. Continued, annual analysis of the MCN data should be able to assess the temporal stability of the hotspots so that—if needed—changes in such prioritized programming can be made; additionally, once adjustment is made for cases that have a reported travel history outside of Zanzibar, hotspots of residual transmission can be identified.

The findings presented here demonstrate that data collected through routing testing of febrile patients for malaria, as well as case follow-up and rACD, can help describe local malaria transmission dynamics at fine spatial scales. The challenge remains to develop programmatically affordable and scalable approaches using routine data that allow for the identification of local spatial heterogeneity to consider more tailored and targeted prevention and control efforts.

## Data Availability

All data produced in the present study are available upon reasonable request to the authors.

## Acknowledgements

Financial support for this study was provided by the U.S. President’s Malaria Initiative through the U.S. Agency for International Development *Okoa Maisha Dhibiti Malaria* Activity (Cooperative Agreement Number: 72062118CA-00002). The findings and conclusions in the manuscript are those of the authors and do not necessarily reflect the views of the President’s Malaria Initiative, the U.S. Agency for International Development, the U.S. Centers for Disease Control and Prevention, or other employing organizations or sources of funding.

## Authors’ contributions

DB, SL, FBA, CK, NS, ER, EE, and RR conceptualized and designed the study. SL, HRM, AA, and JJJ managed and curated the data. DB conducted the data analyses. SL, FAB, SSN, EE and RR supervised the execution of the study. DB, ER and RR drafted the manuscript; all authors read and approved the final version of the manuscript.

